# RAGCBPNet: An Efficient Feature Fusion Framework for Wearable Cuffless Blood Pressure Monitoring and Long-term Validation in Real-world Settings

**DOI:** 10.1101/2025.10.30.25339198

**Authors:** Hongda Huang, Xiao Peng, Xiaoyu Li, Shuailong Tang, Guangpu Zhu, Xiajiao Yang, Hongwei Li, Yelei Li, Yali Zheng

**Affiliations:** Department of Biomedical Engineering, College of Health Science and Environmental Engineering, Shenzhen Technology University, Shenzhen, China; OPPO Health Lab, Shenzhen, China; Tibet Branch Hospital of West China Hospital, Sichuan University, Sichuan, China

## Abstract

Wearable and cuffless blood pressure (BP) monitoring hold great promise for preventive hypertension management, yet few studies have been validated under real-world and long-term conditions. In this study, we propose RACGBPNet, an efficient yet effective feature fusion framework for cuffless BP estimation and hypertension detection. The model leverages a two-stage fusion strategy: first, handcrafted and deep PPG representations are integrated via a self-attention mechanism to capture dynamic signal interactions; second, a simple gating module adaptively fuses signal features with demographic information. To mitigate severe class imbalance in hypertension detection, we further introduce a supervised contrastive learning scheme with a pretrained regression encoder. RACGBPNet was evaluated on two large-scale datasets, including a long-term smartwatch dataset spanning 30+ days in free-living conditions. On the public Aurora-BP dataset, RACGBPNet achieved state-of-the-art performance with mean absolute errors (MAEs) of 7.08 and 4.90 mmHg for systolic and diastolic BP, and Area Under the Receiver Operating Characteristic Curve (AUROC) of 0.8627 and 0.8301 for systolic and diastolic hypertension detection under highly imbalanced data. Long-term validation confirmed that baseline calibration was sufficient to maintain compliance with AAMI standards for SBP up to 14 days and for DBP across the entire tracking period, striking a practical balance between accuracy and usability. However, the model exhibited significantly reduced variability compared with reference BP, indicating challenges in capturing abrupt BP changes. Overall, this work demonstrates the feasibility of long-term smartwatch-based BP monitoring and offers new insights into real-world deployment settings, particularly regarding calibration frequency and cross-dataset generalization.

**Author summary:** Hypertension is one of the most important risk factors for cardiovascular diseases. Wearable devices based on photoplethysmography (PPG) have been increasingly adopted by the general population for daily health monitoring, offering great platform for early detection, prevention and management of hypertension. However, very few studies have examined whether these devices can accurately monitor blood pressure (BP) and detect hypertension in real-life settings for extended periods. We developed a novel framework, RACGBPNet, which efficiently fuses PPG features with personal information for accurate BP estimation, and addresses data imbalance to enhance hypertension detection. Validation on a new smartwatch dataset including 136 subjects, 24,910 recordings collected over more than 30 days in real-life scenarios showed high accuracy over weeks with simple baseline calibration. Our findings also provide valuable insights into calibration frequency, paving the way for reliable, real-world deployment of cuffless BP monitoring techniques.

## 1. Introduction

Hypertension is a leading risk factor for cardiovascular diseases (CVDs), such as myocardial infarction and stroke, which remain the primary causes of mortality worldwide [1]. Nearly half of adults aged 30 to 79 with hypertension are unaware of their condition [2]. Current diagnosis of hypertension typically relies on repeated office blood pressure (BP) measurements [3] or 24-hour ambulatory BP monitoring (ABPM) [4]. These methods are cuff-based, and often disrupt daily activities and sleep, thereby limiting their suitability for continuous and long-term monitoring [5].

To overcome the limitations of cuff-based techniques, cuffless BP monitoring methods have emerged as a major research and industrial focus [6]. BP can be estimated from physiological signals such as photoplethysmography (PPG), electrocardiography (ECG), or pulse pressure waveform (PPW) [7]. Among these, PPG is especially attractive due to its low cost, ease of acquisition and seamless integration into consumer wearables. Over the past decade, numerous cuffless BP estimation models have been developed based on PPG signals, which broadly fall into two categories: mechanism-driven and data-driven models. Mechanism-driven models are grounded in well-established biophysical foundations including Moens-Kortweg and Bramwell-Hill equations [8], but needs individual calibration and struggle to capture arterial wall dynamics in real-world conditions [9, 10]. Data-driven models, in contrast, leverage machine learning to extract feature from physiological signals, yielding improved accuracy [11-14]. The advent of deep learning has further boosted performance by automatic representation learning [15]. Commonly adopted models include convolutional neural networks (CNNs) [16], long short-term memory (LSTM) networks to capture temporal dependencies [17], and hybrid CNN-LSTM architectures to combine their strengths [18]. More recently, Transformer architectures with self-attention mechanisms have demonstrated strong capability in modeling long-range dependencies and have also been applied to cuffless BP estimation [19-21]. Further efforts have explored fusing handcrafted features, deep representations and demographic features to enhance estimation accuracy [22-24].

Despite these advances, several challenges remain unresolved. First, the computational efficiency of complex deep models such as Transformers has not been systematically justified, which may pose great challenges in their applicability to resource-constrained wearable devices. Second, for hypertension detection, class imbalance particularly the underrepresentation of Stage II hypertension, may pose challenges to accurate classification. Furthermore, very few works have investigated long-term validation in real-world settings [25], particularly regarding model generalizability across datasets and calibration requirements over extended monitoring periods.

To address these gaps, we propose RAGCBPNet (ResNet enhanced by Attention, Gating and Contrastive learning for BP estimation), an efficient feature fusion framework for wearable, cuffless BP monitoring and hypertension detection. Our framework introduces a strategic fusion of high-dimensional PPG signal features and demographic information using self-attention and gating mechanisms, alongside regression-based pre-training combing supervised contrastive learning to mitigate class imbalance for accurate hypertension detection. We further evaluate the model on both large-scale public and long-term real-world datasets, providing new insights into calibration frequency and cross-dataset generalization for practical deployment. The main contributions of this study are as follows:

1. *Efficient feature fusion*: We designed a simple yet effective model (RACGBPNet) that integrates self-attention and gating mechanisms to strategically fuse handcrafted features, deep representations and demographic information, achieving state-of-the-art accuracy on the Microsoft Aurora-BP dataset.
2. *Improved hypertension detection*: We introduced a supervised contrastive learning framework with a class-balanced sampling strategy, complemented by regression-based pretraining to address class imbalance issue, leading to more discriminative feature representations.
3. *Real-world long-term validation*: We validated RAGCBPNet on a smartwatch dataset collected over 30+ days under free-living conditions, demonstrating feasibility for long-term monitoring and providing insights into calibration frequency and cross-dataset generalization for real-world deployment.

## 2. Related Work

### 2.1 Datasets

Most existing studies were validated on small datasets. For example, one dataset included 85 participants (mean age 49.9 ± 9.0 years), with each subject undergoing BP measurements in a resting supine position [26]. Another study recruited 89 middle-aged and elderly subjects (mean age 49.5 ± 7.9 years) and collected 3,104 recordings across multiple days, with simultaneous ECG and radial-artery PPG acquisition after cuff-based BP measurements [27]. The PPG-BP dataset containing 657 data records from 219 subjects was also widely used [28]. In addition, several clinical ward-based datasets have been broadly adopted, including MIMIC-III from ICU patients [29], VitalDB from intraoperative patients [30], and PulseDB which combines multiple sources and specifically curated for cuffless BP research [31]. More recent work has shifted toward large-scale, heterogeneous datasets to improve robustness and generalizability. The Microsoft Aurora-BP dataset includes both static and 24-hour ambulatory measurements from 1,125 subjects [32], while the CAS-BP dataset contains recordings from 3,077 subjects over four sessions spanning one month (Day 0, Day 7, Day 14, and Day 21) [33]. Beyond controlled data collection, researchers have also begun to explore real-world, long-term monitoring. For example, a team from Seoul National University conducted an observational study using the Samsung Galaxy Watch, demonstrating the feasibility of wearable cuffless BP monitoring under free-living conditions [25].

### 2.2. Models for BP Regression

Mechanism-driven approaches are primarily grounded in vascular biomechanics and pulse wave propagation theory. Based on the classical Moens-Korteweg and Hughes equations [10], Poon et al. proposed a pulse transit time (PTT) based model for separately estimating systolic (SBP) and diastolic blood pressure (DBP) [34]. Subsequent studies further incorporated additional physiological parameters. For example, Ding et al. demonstrated that the photoplethysmographic intensity ratio can reflect arterial diameter changes, enabling low-frequency BP tracking with particular sensitivity to DBP [35]. Liu et al. introduced a model incorporating small-artery PTT derived from multi-wavelength PPG, which served as an indicator of peripheral vascular resistance and achieved superior BP tracking compared to conventional PTT-based models [36].

In contrast, data-driven approaches extract informative features directly from physiological signals, enabling them to capture complex nonlinear relationships between input signals and BP. CNNs have been widely applied for automated waveform feature extraction, such as an enhanced ResNet which integrated a multi-scale feature extraction module and channel-attentive residual blocks [37]. Recurrent neural networks, particularly LSTM networks, have been employed to model long-range temporal dependencies [17]. Hybrid CNN-LSTM architectures have also been proposed to jointly exploit spatial and temporal patterns, with attention mechanisms highlighting the most relevant temporal features [18]. Building on these developments, our previous work proposed UTransBPNet, which employed a Transformer backbone and integrated a multi-head cross-attention module to refine both short- and long-range dependencies, achieving superior performance in dynamic conditions [38]. Similarly, Liu et al. designed a manual feature-guided CNN-Transformer model that incorporated handcrafted and learned representations through a dedicated fusion module [23]. Beyond purely data-driven learning, some studies have explored physics-informed neural networks to embed Taylor approximation of gradually varying relationship between cardiovascular variables into the training process, thereby reducing the reliance on large amount of ground truth data. However, such methods still require subject-specific calibration [39].

### 2.3. Models for BP Classification

Beyond continuous BP estimation, hypertension detection is also of substantial clinical value, enabling earlier diagnosis and management. Traditional machine learning approaches have been widely explored. For example, Nour et al. applied a decision tree classifier with handcrafted features on the PPG-BP dataset [22], while Evdochimet et al. adopted both decision trees and support vector machines for BP classification using the MIMIC-III dataset [23]. Gupta et al. combined CNNs for spatial feature extraction with LSTMs for temporal modeling, achieving improved performance on the PPG-BP dataset [24]. Similarly, Sayed et al. introduced ExHyptNet, a hybrid framework that leverages recurrent graph networks and EfficientNetB3 for PPG feature extraction, together with ensemble classifiers such as XGBoost and extremely randomized trees [40]. Notably, a recent large-scale study involving over 32,000 participants and more than 660,000 paired PPG and cuff-based BP records demonstrated that PPG alone provides strong discriminatory power for hypertension detection with XGBoost [40].

## 3. Materials and Methods

### 3.1. Datasets

The proposed model was developed and evaluated using two datasets. The first is the Microsoft Aurora-BP dataset [32] containing both static and ambulatory measurements. For this study, only the static measurements were used, with reference BP obtained via an auscultatory device (Diagnostix™ 700/703, American Diagnostic Co., USA). To ensure consistency with the second dataset, only PPG signals were retained for BP modeling. A rigorous preprocessing pipeline was applied: 427 recordings with missing entries were excluded, followed by the removal of 1,053 recordings with a signal quality score (provided with the dataset) below 0.5. Additionally, 94 recordings with BP beyond physiological ranges (SBP ≥ 220 mmHg or DBP ≤ 30 mmHg) were discarded. After preprocessing, the final dataset consisted of 9,433 recordings from 626 subjects.

The OPPO-BP dataset was collected in accordance with a standardized home BP monitoring protocol, which was approved by the Ethics Committee of the Tibet Branch of the West China Hospital of Sichuan University, and written informed consent was obtained from all participants prior to enrollment. PPG signals were acquired using OPPO smartwatches, with each recording lasting approximately 60 seconds at a sampling rate of 250 Hz. Reference BP was measured using an Omron electronic sphygmomanometer. Participants were instructed to perform at least three BP measurements during each of three daily periods (morning, noon, and evening), with a minimum interval of one minute between successive measurements. To ensure measurement stability, additional readings were taken if the difference between consecutive measurements exceeded 12 mmHg for SBP or 8 mmHg for DBP. The duration of data collection varied across participants, ranging from several days up to approximately 40 days.

Table 1 summarizes the demographic and statistical characteristics of the two datasets. The Aurora-BP dataset includes a relatively older population, whereas the OPPO-BP dataset contains a substantially larger number of recordings, primarily due to the extended collection period. Both datasets exhibited marked class imbalance, with hypertension defined as SBP > 140 mmHg or DBP > 90 mmHg [41]. Fig. 1 illustrates the BP distribution of the two datasets, and Fig. 2 presents the distribution of recording durations for the OPPO-BP dataset. Notably, most participants in the OPPO-BP cohort contributed data for approximately 35 days.

**Fig. 1.**
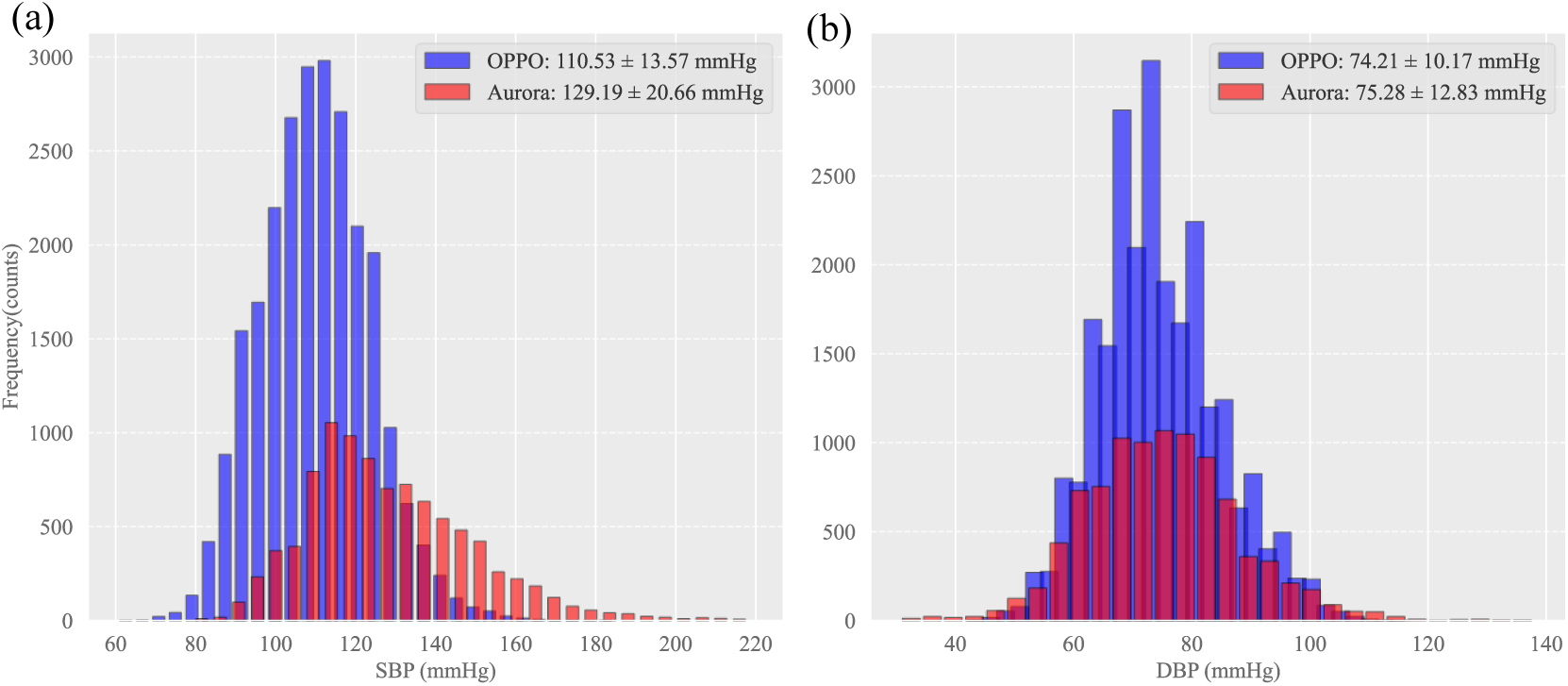
BP distribution of Aurora-BP and OPPO-BP datasets. (a) SBP; (b) DBP.

**Fig. 2.**
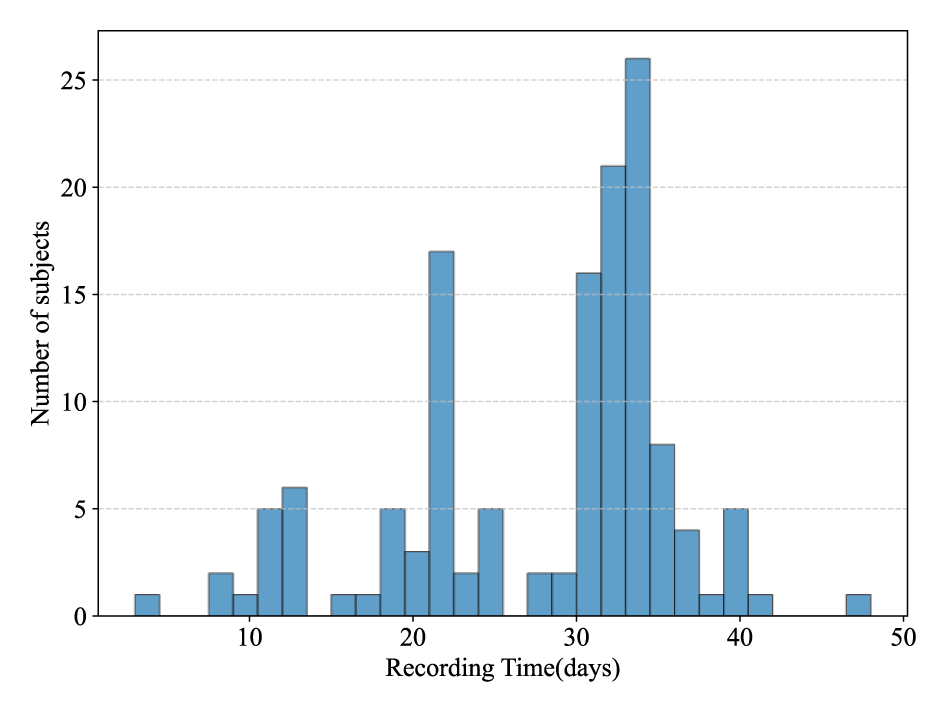
Distribution of the recording durations for the OPPO-BP dataset.

**Table 1.**
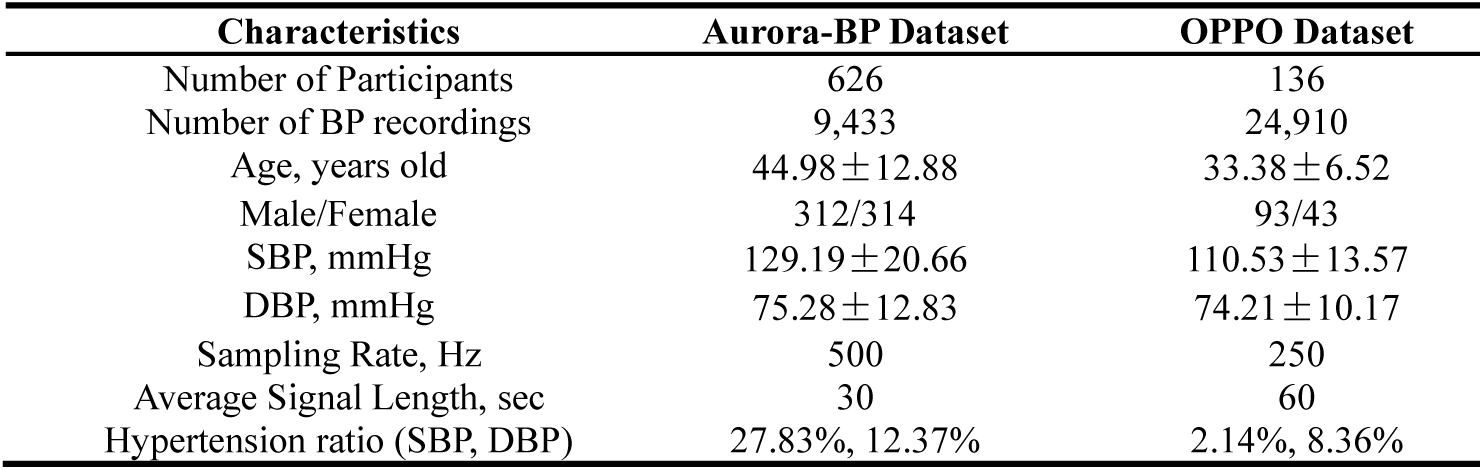
Demographics and Statistics of Aurora-BP and OPPO-BP Datasets.

| Characteristics | Aurora-BP Dataset | OPPO Dataset |
| --- | --- | --- |
| Number of Participants | 626 | 136 |
| Number of BP recordings | 9,433 | 24,910 |
| Age, years old | $44.98 \pm 12.88$ | $33.38 \pm 6.52$ |
| Male/Female | 312/314 | 93/43 |
| SBP, mmHg | $129.19 \pm 20.66$ | $110.53 \pm 13.57$ |
| DBP, mmHg | $75.28 \pm 12.83$ | $74.21 \pm 10.17$ |
| Sampling Rate, Hz | 500 | 250 |
| Average Signal Length, sec | 30 | 60 |
| Hypertension ratio (SBP, DBP) | 27.83%, 12.37% | 2.14%, 8.36% |

### 3.2. Data Preprocessing

Raw PPG signals were first preprocessed using a Butterworth band-pass filter (0.25-10 Hz) to reduce baseline drift and high-frequency noise. Baseline correction was then applied using the adaptive iteratively reweighted penalized least squares (airPLS) algorithm [42]. A total of 476 handcrafted features were extracted from the preprocessed PPG signals following the method proposed by Cisnal et al. [43]. This feature set included 404 time-domain features, 58 frequency-domain features, and several statistical descriptors. The preprocessed PPG signals were then downsampled to 125 Hz and normalized using min–max scaling. The normalized signals, together with their first- and second-order derivatives, were subsequently used as inputs to the model for deep feature learning.

### 3.3. Modeling

The architecture of the proposed RACGBPNet model is illustrated in Fig. 3. It integrates demographic information, deep learning features, and handcrafted features with a two-stage strategy to enhance BP estimation. In the first stage, deep features learned from PPG signals were fused with handcrafted features using a self-attention module, which enables dynamic interactions across multi-source signal representations. In the 2^nd^ stage, a gating module performs a simpler yet effective fusion by adaptively selecting salient features from the abstract signal representations and demographic features. This hierarchical fusion enables the model to capture rich, fine-grained dependencies within high-dimensional signal representations, while maintaining efficiency when integrating contextual demographic information. The details of each component are described below.

**Fig. 3.**
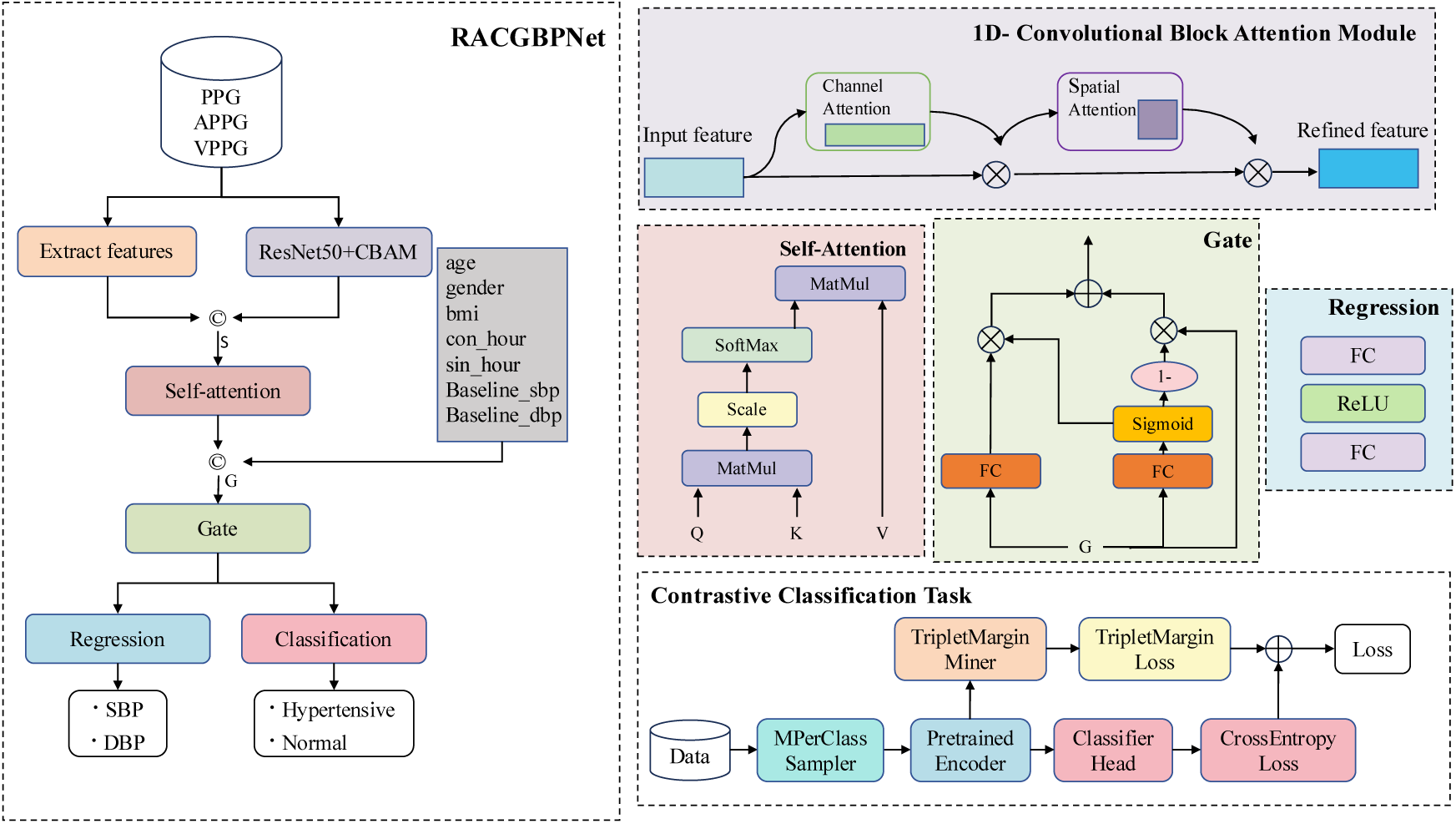
The proposed architecture of RACGBPNet.

#### 3.3.1 Deep Feature Extraction Module

ResNet50, a widely used deep convolutional neural network with residual connections, was adopted as the backbone of the deep feature extractor for learning hierarchical representations from complex PPG signals. Such deep architectures are essential for capturing subtle morphological variations and temporal dependencies in PPG signals that cannot be effectively modeled by handcrafted features alone. To further refine representation quality, a Convolutional Block Attention Module (CBAM) was integrated into ResNet50, enabling the network to adaptively refine feature maps by emphasizing informative regions and suppressing irrelevant ones, without significantly increasing computational complexity.

#### 3.3.2 Feature fusion: Self-Attention and Gating Modules

ResNet50, a widely used deep convolutional neural network with residual connections, was adopted as the backbone of the deep feature extractor for learning hierarchical representations from complex PPG signals. Such deep architectures are essential for capturing subtle morphological variations and temporal dependencies in PPG signals that cannot be effectively modeled by handcrafted features alone. To further refine representation quality, a Convolutional Block Attention Module (CBAM) was integrated into ResNet50, enabling the network to adaptively refine feature maps by emphasizing informative regions and suppressing irrelevant ones, without significantly increasing computational complexity.

While the self-attention module captures fine-grained dependencies between deep and handcrafted features, the gating module provides a subsequent adaptive feature selection. It filters the fused signal representations together with baseline demographic and other variables (age, gender, height, weight, BMI, baseline SBP, baseline DBP, and circadian encodings of acquisition hour). Inspired by the design principles of the Gated Recurrent Unit (GRU) [44], the gating mechanism dynamically regulates the contribution of each input feature, enabling the model to retain salient information while maintaining computational efficiency. The mathematical formulation is given in Equation (1),

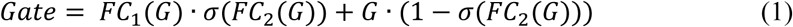

where *G* denotes the input feature vector, *FC* represent fully connected layers, and *σ* denotes the sigmoid activation function.

#### 3.3.3 Projection Heads for Regression and Classification

The regression head comprises two fully connected layers, with a ReLU activation function in between to introduce non-linearity. The features obtained with the 2-stage fusion strategy were concatenated with the baseline SBP/DBP values were fed into the regression head to further enhance the contribution of baseline BP.

For hypertension detection, several strategies were employed to mitigate the class imbalance issue. First, a sampling strategy named MPerClassSampler, was utilized to ensure each batch contains an equal number of samples from each class. Second, to enhance feature separability and accelerate convergence, a supervised contrastive learning scheme based on TripletMarginLoss was introduced [45], which encourages embeddings of the same class to cluster while pushing embeddings of different classes apart.

Specifically, for an anchor sample *a*_*i*_, a positive sample *p*_*i*_ (from the same class), and a negative sample *n*_*i*_ (from a different class), the TripletMarginLoss is formulated as:

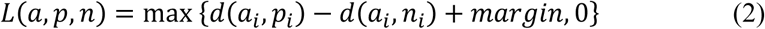

where the distance function is the Euclidean distance:

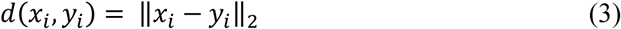

The margin was set to 0.2, and hard triplet mining was applied, selecting only the most challenging triplets within each batch (i.e., those closest to violating the margin constraint). Finally, to further enhance discriminative power, the encoder (all layers preceding the projection head) was initialized with weights from a pretrained regression model.

### 3.4 Model Training and Evaluation

A five-fold cross validation scheme was applied within each dataset, using subject-wise partitioning to prevent data leakage. For the regression task, the loss function was the Mean Absolute Error (MAE), whereas for the classification task, the cross-entropy loss was used. In the contrastive learning–based classification setting, both cross-entropy loss and TripletMarginLoss were jointly optimized. The total loss is formulated as:

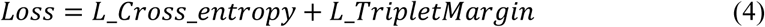

All models were trained using the Adam optimizer for 200 epochs with a batch size of 128. For both regression and standard classification tasks, the learning rate and weight decay were set to 0.001. In the contrastive classification task, a two-tier learning rate strategy was adopted: the encoder was trained with a reduced learning rate and weight decay of 0.0001, while the classification head retained 0.001. To mitigate overfitting, early stopping with a patience of 20 was applied based on validation performance, and the model with the highest validation accuracy was selected for testing in each fold. All experiments were implemented in PyTorch and executed on an NVIDIA Tesla V100 PCIe GPU (32 GB RAM).

#### Evaluation Metrics

For regression, the performance was assessed following standard international standards using Mean Error (ME), Mean Absolute Difference (MAD), Standard Deviation (SD) of the difference between estimated and reference BP values. For classification, the following metrics were reported: Accuracy, Precision, Recall, F1 Score, Area Under the Receiver Operating Characteristic Curve (AUROC), and Area Under the Precision-Recall Curve (AUPRC).

#### Cross-dataset Validation

To assess generalization, cross-dataset evaluation was conducted by training on the Aurora-BP dataset and testing on the OPPO-BP dataset. The OPPO-BP test set was normalized using the same scheme as the training data. For model selection, 10% of OPPO-BP was randomly sampled while ensuring subject independence from the remaining 90% test set. The Aurora-BP model with the best performance on this 10% subset was used for the final cross-dataset evaluation.

## 4. Results

### 4.1. Comparison with Baseline Models

**Table 2** presents the performance comparison between RACGBPNet and several regression models evaluated on the Aurora-BP dataset. RACGBPNet achieved the best overall performance, with MAEs of 7.08 mmHg for SBP and 4.90 mmHg for DBP. These results represent statistically significant improvements over traditional machine learning models. Compared with the second-best model, ResNet50, RACGBPNet achieved an additional performance gain of 0.39 mmHg for SBP. For fairness of comparison, the state-of-the-art HGCTNet model [21] was retrained in this study using PPG, VPPG, and APPG signals as inputs.

**Table 2.** Performance comparison of RACGBPNet with baseline models for regression tasks.

| Model | SBP |  | DBP |  |
| --- | --- | --- | --- | --- |
| | MAE (mmHg) | ME ± STD (mmHg) | MAE (mmHg) | ME ± STD (mmHg) |
| Ridge [32] | - | 0.42 ± 10.10 | - | 0.55 ± 7.66 |
| Decision Tree | 8.48* | -0.14 ± 12.42 | 5.78* | 0.19 ± 8.04 |
| Gradient Boost [43] | 8.49 | 0.63 ± 11.73 | <u>5.03</u> | 0.01 ± 6.58 |
| Adaboost | 7.65* | -0.47 ± 10.78 | 5.30* | 0.47 ± 7.20 |
| Resnet50 [46] | <u>7.47</u> | 0.85 ± 11.05 | 5.13* | -0.18 ± 7.12 |
| Vggnet16 [27] | 7.83 | 1.67 ± 12.02 | 5.25* | -0.74 ± 7.50 |
| HGCTNet [21] | 7.61 | 1.24 ± 11.52 | 5.21 | -0.90 ± 7.43 |
| RACGBPNet | **7.08** | 0.96 ± 10.29 | **4.90** | -0.46 ± 6.78 |

(Note: \* indicates a significant difference ( $p < 0.05$ ) in MAE between RACGBPNet and other models under Mann-Whitney U test)

Fig. 4 and Fig. 5 visualize the agreement between the estimation by RACGBPNet and the reference through Bland-Altman and correlation plots, respectively. The correlation plots demonstrate strong linear agreement, with correlation coefficients exceeding 0.80 in all cases. In the Bland-Altman plots, the vast majority of data points fall within or close to the 95% limits of agreement, indicating strong consistency between estimated and reference BP values.

**Fig. 4.**
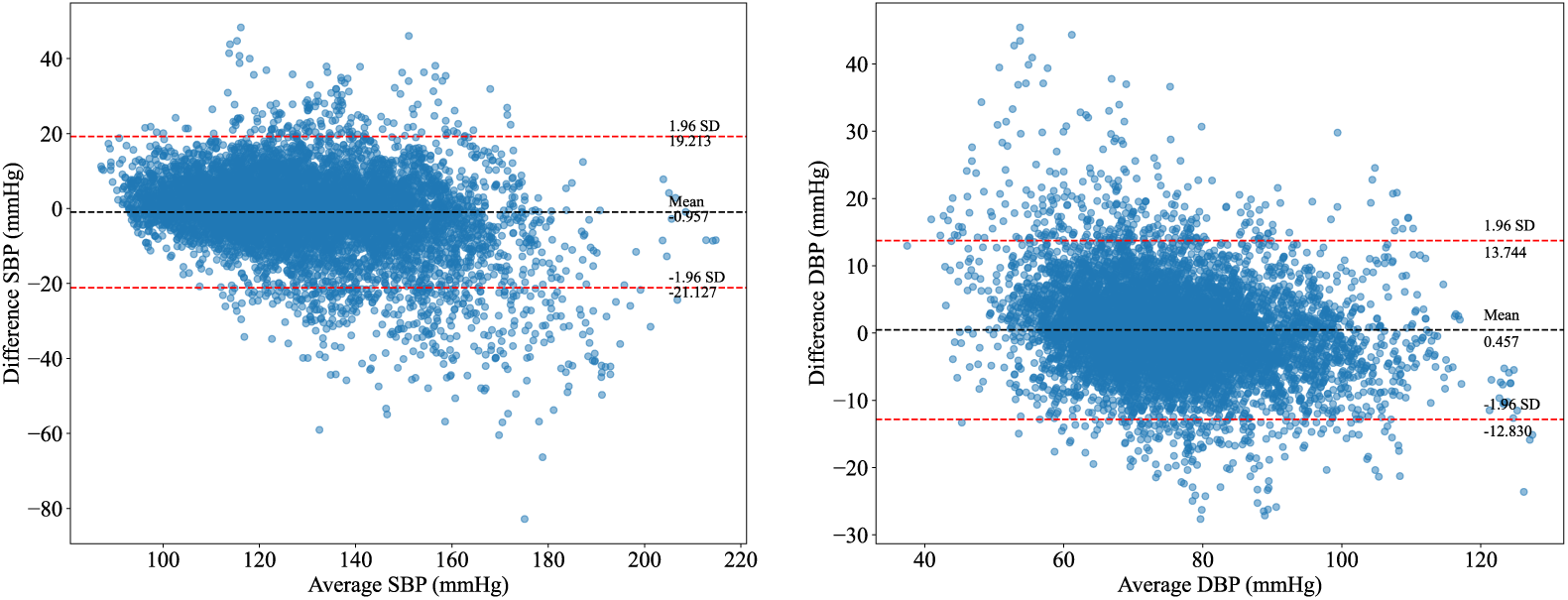
B and-A tman p ots for the proposed RACGBPNet. (a) SBP; (b) DBP.

**Fig. 5.**
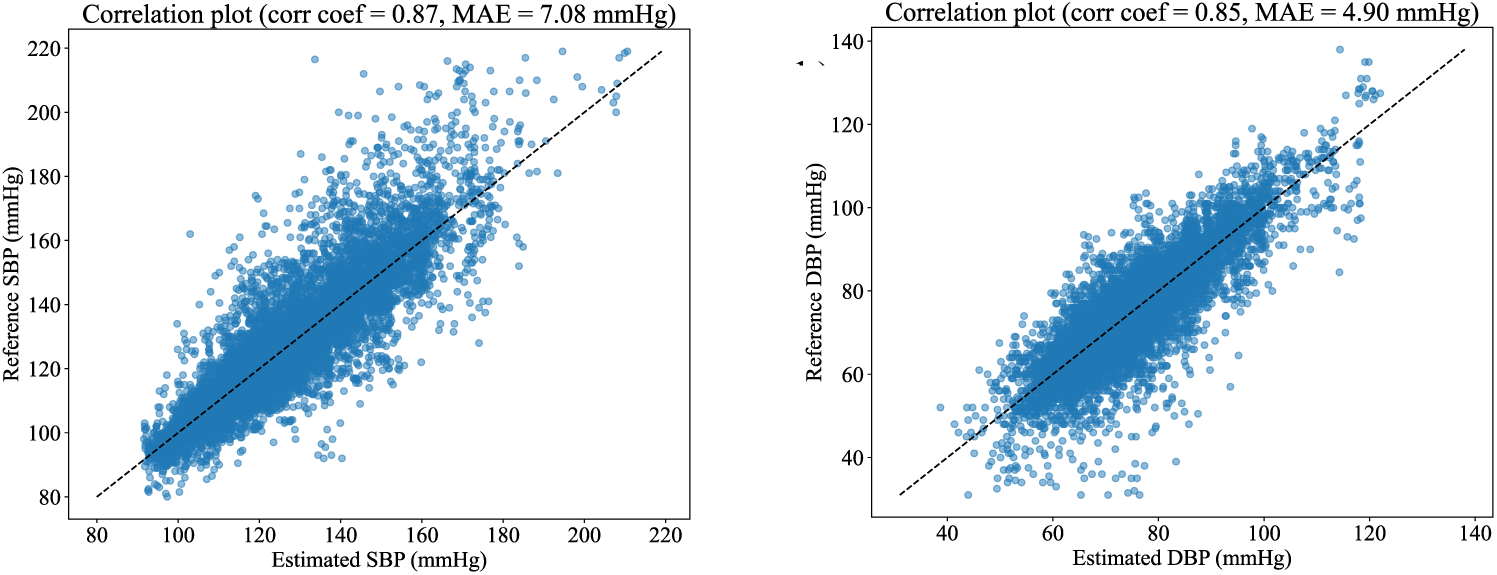
Corre ation p ots for the proposed RACGBPNet. (a) SBP; (b) DBP.

Table 3 summarizes the performance of the proposed RACGBPNet and several baseline models for hypertension detection. The proposed model achieved the accuracy of 89.52% and 94.42% for systolic and diastolic hypertension detection respectively, presenting a slight improvement of 1.17% and 1.04% compared to the best baseline model. In addition, the proposed model outperformed all baseline methods in terms of AUPRC, AUROC and F1 score, indicating its superior performance in class discrimination under imbalance condition.

**Table 3.**
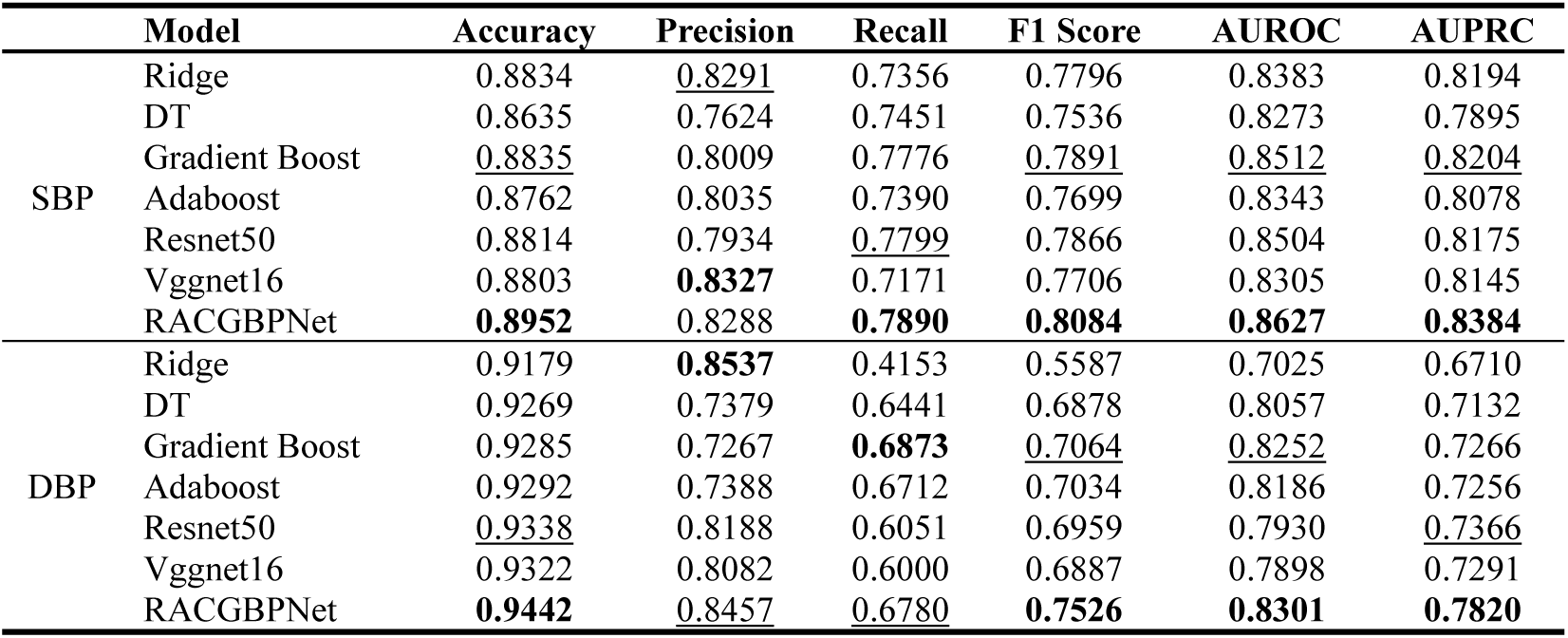
Performance comparison of RACGBPNet with baseline models for classification task.

### 4.2. Ablation Results on Aurora-BP dataset

Table 4 and Table 5 present the ablation results for regression and classification, respectively. The results suggest that every component contributes to the improvement of overall accuracy. A comparison between self-attention (SA) and multi-head attention (MHA) indicates that, the latter brought marginal performance gain, suggesting that a relatively simple self-attention mechanism is sufficient to capture dynamic interactions between handcrafted and deep features. In contrast, replacing ResNet50 with the CNN architecture used in HGCTNet [21] led to a marked performance drop, highlighting the value of a strong deep feature extractor at the cost of higher computational load and model size. Introducing SA+Gate further improved the MAE from 7.47 to 7.08 mmHg with minimal computational and memory overhead, demonstrating the efficacy and efficiency of the proposed fusion strategy. On the other hand, compared to the classification model trained with a naïve strategy, incorporating supervised contrastive learning together with a pretrained regression encoder yielded the highest accuracy. The performance gain was particularly pronounced for DBP classification, likely due to the more severe class imbalance observed in DBP categories.

**Table 4.**
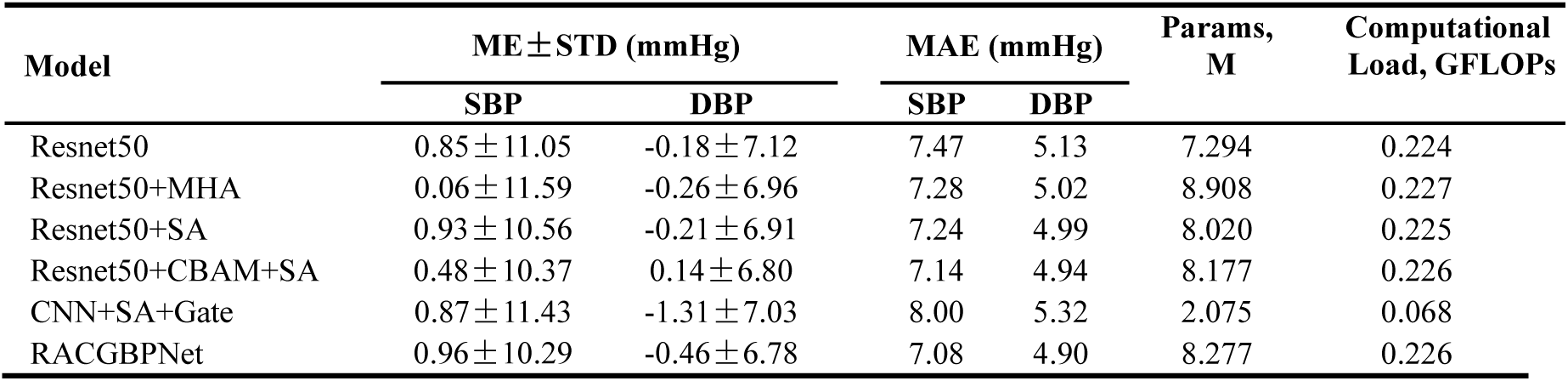
Ablation results for BP regression.

| Model | ME±STD (mmHg) | | MAE (mmHg) | | Params, M | Computational Load, GFLOPs |
| --- | --- | --- | --- | --- | --- | --- |
|  | SBP | DBP | SBP | DBP |  |  |
| Resnet50 | 0.85±11.05 | -0.18±7.12 | 7.47 | 5.13 | 7.294 | 0.224 |
| Resnet50+MHA | 0.06±11.59 | -0.26±6.96 | 7.28 | 5.02 | 8.908 | 0.227 |
| Resnet50+SA | 0.93±10.56 | -0.21±6.91 | 7.24 | 4.99 | 8.020 | 0.225 |
| Resnet50+CBAM+SA | 0.48±10.37 | 0.14±6.80 | 7.14 | 4.94 | 8.177 | 0.226 |
| CNN+SA+Gate | 0.87±11.43 | -1.31±7.03 | 8.00 | 5.32 | 2.075 | 0.068 |
| RACGBPNet | 0.96±10.29 | -0.46±6.78 | 7.08 | 4.90 | 8.277 | 0.226 |

**Table 5.** Ablation results for hypertension detection.

| Model | Accuracy | Precision | Recall | F1 Score | AUROC | AUPRC |
| --- | --- | --- | --- | --- | --- | --- |
| *SBP* |  |  |  |  |  |  |
| Classification | 0.8885 | <u>0.8289</u> | 0.7587 | 0.7923 | 0.8489 | 0.8276 |
| ~ + Pretrain | 0.8896 | **0.8372** | 0.7526 | 0.7927 | 0.8478 | 0.8296 |
| ~ + Contrastive | <u>0.8920</u> | 0.8175 | **0.7912** | 0.8042 | <u>0.8612</u> | <u>0.8336</u> |
| ~ + Contrastive+ Pretrain | **0.8952** | 0.8288 | <u>0.7890</u> | **0.8084** | **0.8627** | **0.8384** |
| *DBP* |  |  |  |  |  |  |
| Classification | 0.9372 | 0.8303 | 0.6263 | 0.7140 | 0.8040 | 0.7517 |
| ~ + Pretrain | 0.9384 | 0.8196 | 0.6508 | 0.7256 | 0.8152 | 0.7571 |
| ~ + Contrastive | <u>0.9424</u> | <u>0.8328</u> | <u>0.6754</u> | <u>0.7459</u> | <u>0.8280</u> | <u>0.7744</u> |
| ~ + Contrastive+ Pretrain | **0.9442** | **0.8457** | **0.6780** | **0.7526** | **0.8301** | **0.7820** |

### 4.3. Individual BP Trends

Fig. 6 illustrates the trends of estimated and reference BP values for several individuals across both datasets, with background colors distinguishing different individuals. As observed, the variability of reference BP is significantly greater than that of the estimated values (Wilcoxon signed-rank test, *p* < 0.00). The model is able to follow the overall trend but struggles to capture abrupt fluctuations. Fig. 7 demonstrated the distribution of individual deviation level from baseline (grouped as 10 mmHg bins) and their relationship with model performance for Aurora-BP dataset. The results indicate that MAE increased consistently with deviation level, suggesting that larger deviations together with the scarcity in higher-deviation bins make accurate estimation more challenging.

**Fig. 6.**
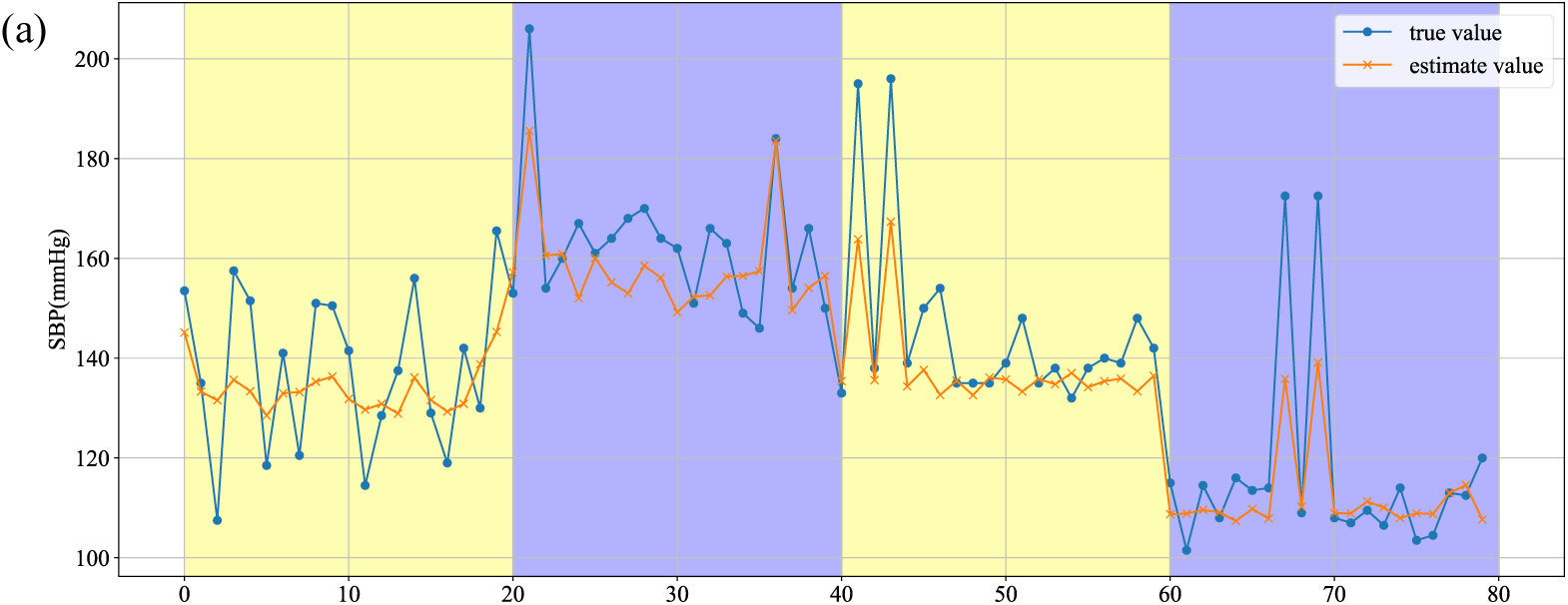
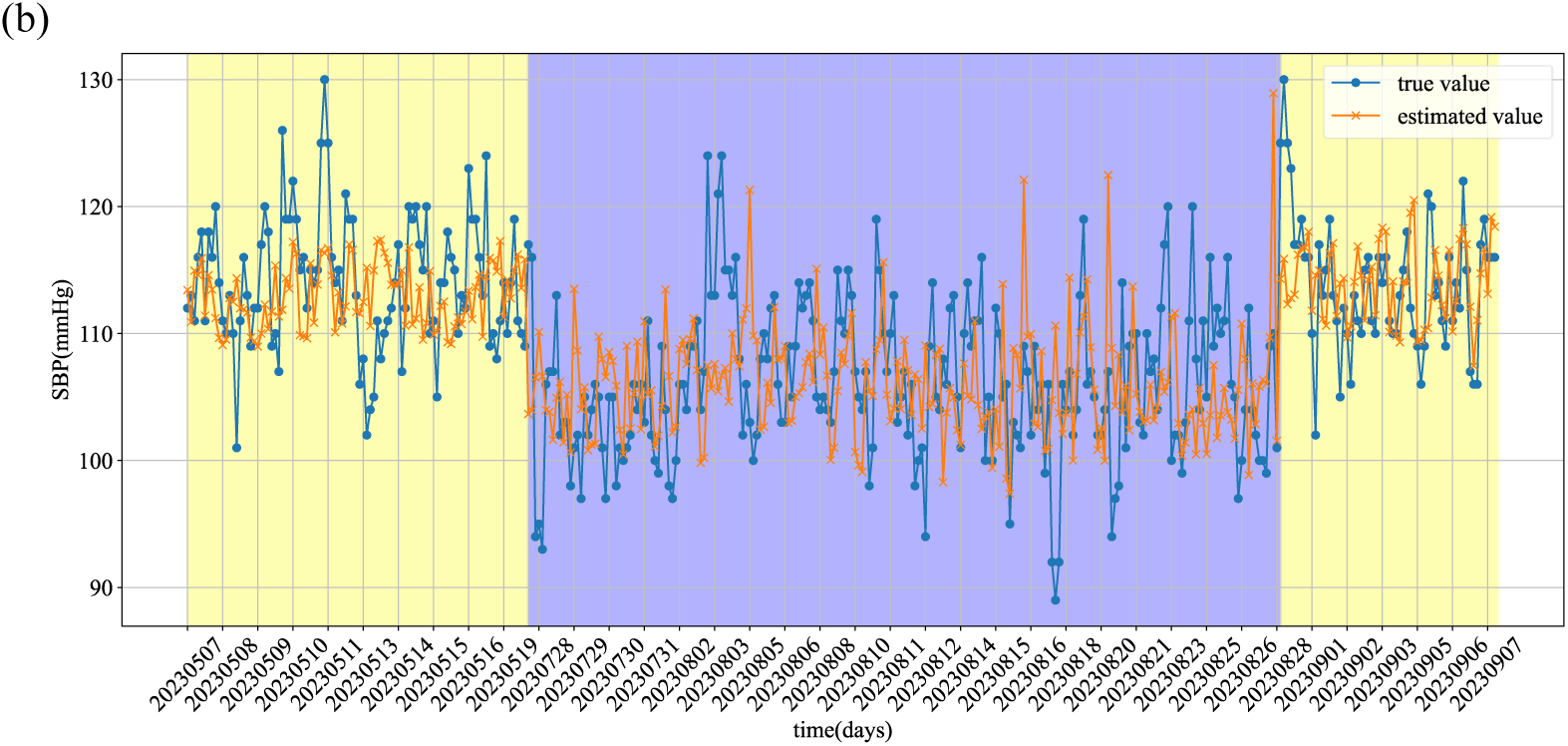
The trends of reference and estimated BP for some indi idua s. (a) Aurora-BP; (b) OPPO-BP.

**Fig. 7.**
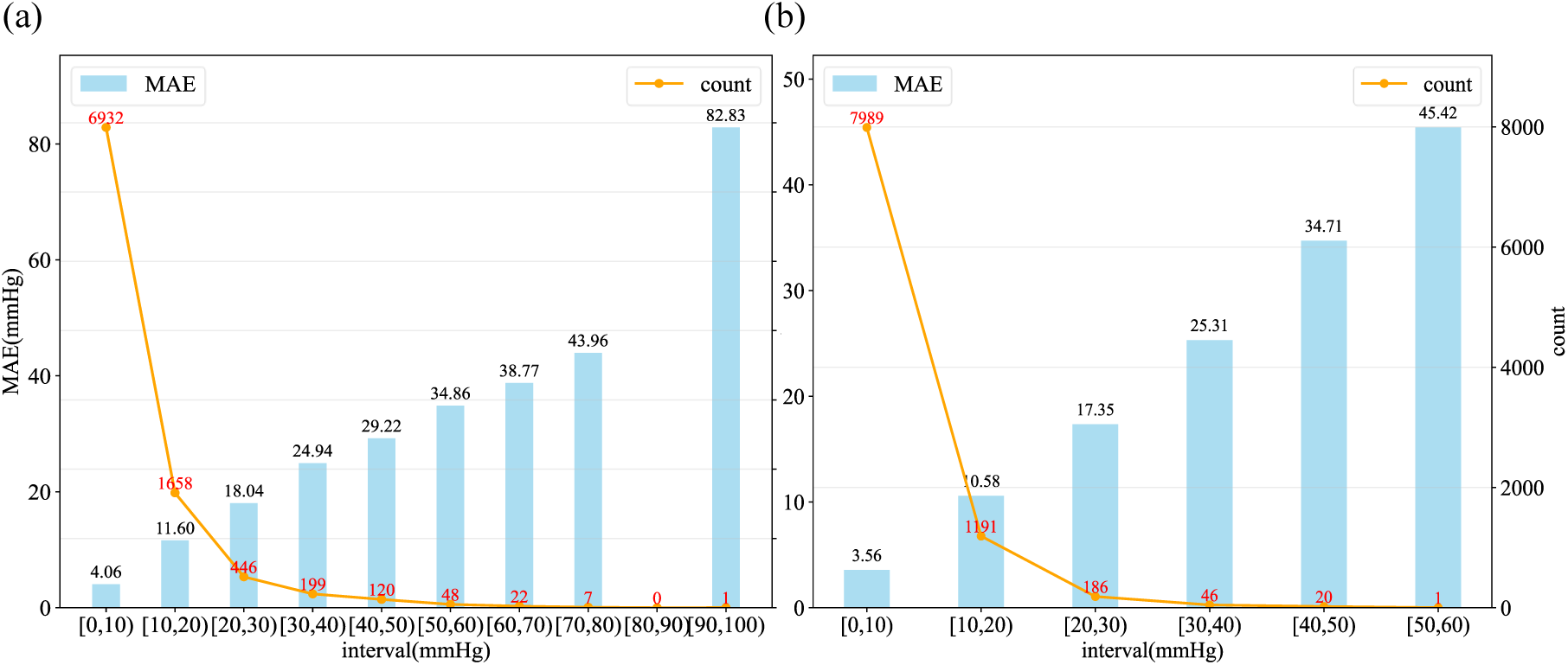
The distribution of the BP de iation e e s from indi idua base ine and the AE at each e e for Aurora-BP dataset. (a) SBP; (b) DBP.

### 4.4. Long-Term Tracking Capability and Influence of Calibration Frequency

Fig. 8 demonstrates the long-term tracking capability of the proposed model in the OPPO-BP dataset. The ME remained below 5 mmHg across the monitoring period, while both MAE and STD showed a gradual increase of less than 2 mmHg for SBP, suggesting a slow performance decline over time. Specifically, during the first 14 days, the SBP accuracy met the AAMI standard (i.e., ME ± STD < 5 ± 8 mmHg). Beyond this period, however, the MAE for SBP exceeded the AAMI standard but remained within the IEEE Standard Grade C range (6–7 mmHg) for approximately 30 days. DBP accuracy was maintained throughout the entire monitoring period.

**Fig. 8.**
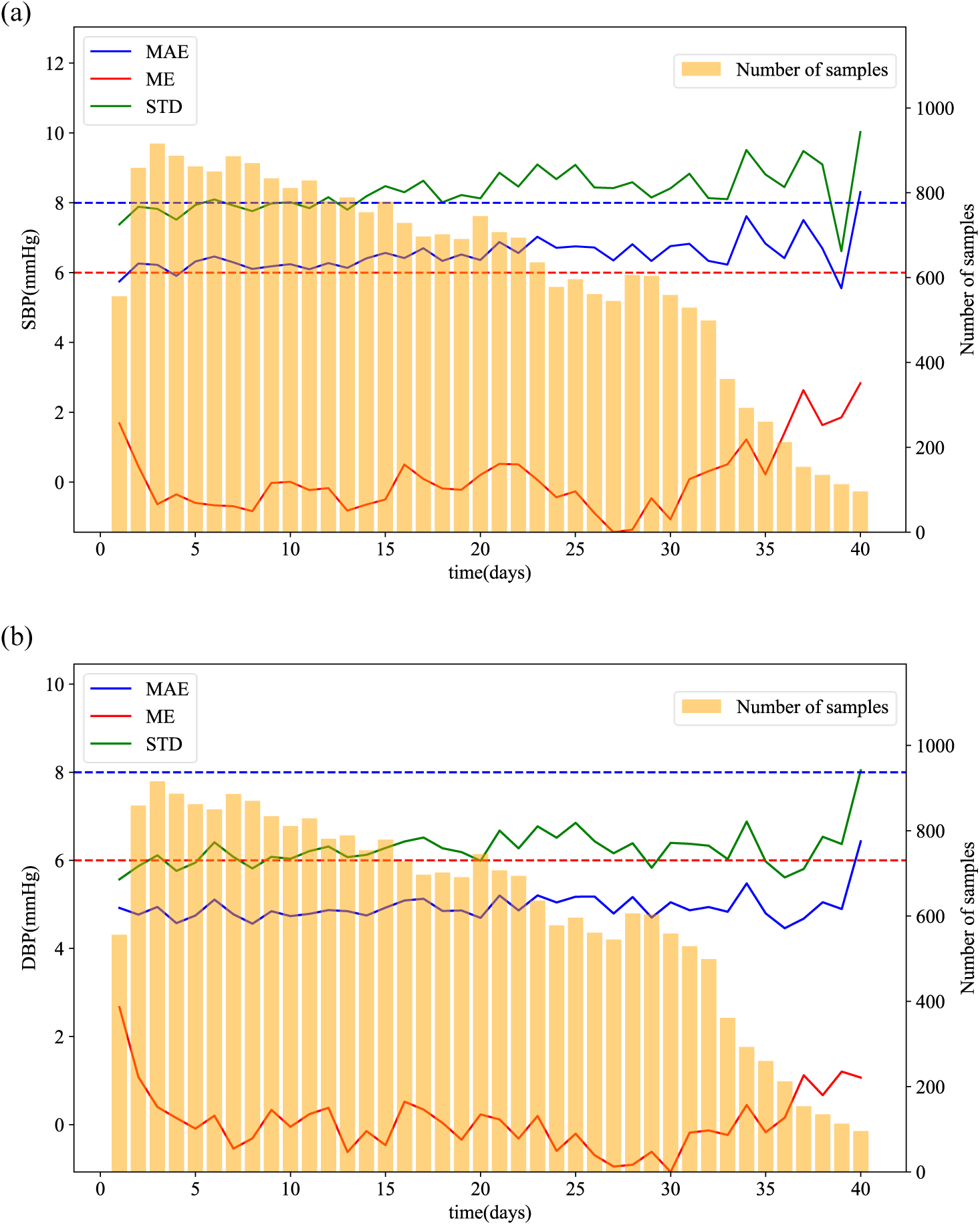
The mode performance o er an extended period of time for OPPO-BP dataset (a) SBP; (b) DBP.

These findings suggest that periodic calibration is necessary to retain accuracy for clinical applicability. Therefore, we further investigated the impact of different calibration intervals on model accuracy as presented in Table 6. Six calibration frequencies were tested: daily, every three days, weekly, every fourteen days, monthly, and a single overall calibration. For each setting, the first BP value within each calibration period was used as the baseline BP. The results showed that, daily calibration reduced the MAE to 5.93 mmHg for SBP and 4.54 mmHg for DBP. As the calibration interval increased, SBP errors gradually rose at an approximate rate of 0.016 mmHg per day, i.e., (6.42-5.93)/30 mmHg. Calibration interval longer than a week led to a slight but significant decline compared to daily calibration, while DBP errors remained largely unaffected across calibration intervals and consistently within acceptable error thresholds.

**Table 6.** Model performance at different time intervals on OPPO-BP dataset.

| Recalibration interval | SBP |  | DBP |  |
| --- | --- | --- | --- | --- |
| | MAE (mmHg) | ME±STD (mmHg) | MAE (mmHg) | ME±STD (mmHg) |
| 1 Day | **5.93** | 1.97±7.43 | **4.54** | 1.55±5.59 |
| 3 Days | 6.26 | 1.98±7.83 | 4.83 | 1.64±5.97 |
| 7 Days | 6.38* | 1.59±8.07 | 4.97* | 1.32±6.07 |
| 14 Days | 6.40* | 1.10±8.14 | 4.91* | 0.95±6.18 |
| 30 Days | 6.42* | 0.11±8.24 | 4.87* | 0.20±6.21 |
| Baseline | 6.44* | -0.12±8.28 | 4.89* | 0.02±6.24 |

(Note: \* indicates a significant difference ( $p < 0.05$ ) between daily calibration and other calibration settings under Mann-Whitney U test)

### 4.5. Comparison of Two datasets and Cross-Dataset Validation

Table 7 summarizes the results of within-dataset cross-validation and cross-dataset testing. Overall, the OPPO-BP dataset yielded better cross-validation performance than the Aurora-BP dataset. However, when evaluating cross-dataset generalization, the MAE for SBP and DBP increased by 1.54 and 1.58 mmHg, respectively, compared with the OPPO-BP cross-validation results.

**Table 7.** Within-Dataset Cross-Validation and Cross-Dataset Testing on Two Datasets.

|  | SBP |  | DBP |  |
| --- | --- | --- | --- | --- |
| | MAE (mmHg) | ME±STD (mmHg) | MAE (mmHg) | ME±STD (mmHg) |
| OPPO-BP | 6.44 | -0.12±8.28 | 4.89 | 0.02±6.24 |
| Aurora-BP | 7.08 | 0.96±10.29 | 4.90 | -0.46±6.78 |
| Aurora-BP→OPPO | 8.98 | -5.32±10.80 | 6.47 | -3.16±7.71 |

## 5. Discussion

In this study, we proposed a novel deep learning architecture, RACGBPNet, which achieved outstanding performance on the public, large-scale Aurora-BP dataset. Unlike HGCTNet [21], which used a shallower CNN and a complex Transformer-based fusion strategy with handcrafted-guided deep feature learning, RACGBPNet employs a deeper feature extractor (ResNet50) together with an efficient fusion strategy combining self-attention mechanism for high-dimensional signal features and a simple gating mechanism when integrating contextual demographic features. Ablation studies confirmed that ResNet50 plays a central role in capturing rich PPG representations, while the proposed fusion strategy further enhances performance without causing much computational and storage overheads. We further extended RACGBPNet to hypertension detection. By leveraging the pretrained regression encoder together with class-balanced sampling and TripletMarginLoss-based contrastive learning, the model effectively addressed the class imbalance between normotension and hypertension, demonstrating strong potential for real-world clinical applications.

The analysis of BP deviation from individual baselines indicates that, the limited number of samples at high-deviation levels makes it difficult for the model to accurately capture the input-BP relationship when arterial conditions deviate substantially from baseline. This limitation partly explains the reduced variability of estimated BP compared with reference values, and the increased errors when BP largely deviated from individual baseline, as observed in Fig. 6. It also explains the gradual performance decline observed during long-term tracking (Fig. 8), as BP may deviate from its baseline during long-term daily monitoring.

These findings highlight the necessity of calibration to maintain long-term applicability. A previous large-scale feasibility study using the Samsung smartwatch recommended a 15-day calibration interval to mitigate significant performance degradation in both SBP and DBP, though compliance with international standards over the long term was not reported [25]. Our findings (Table 6) suggest that a calibration interval of at least 7 days is necessary to prevent significant performance decline. With a baseline calibration, model accuracy remained largely compliant with AAMI standards for up to 14 days and with IEEE Grade C for SBP within 30 days, while consistently meeting IEEE Grade A for DBP across the entire tracking period, demonstrating a practical balance between accuracy and usability in real-world monitoring.

When comparing the two datasets, the Aurora-BP dataset is characterized by an older age distribution, higher BP levels, and a right-tailed skewed distribution, all of which make accurate BP estimation more challenging than in the OPPO-BP dataset. These findings are consistent with previous studies on performance degradation in hypertensive and elderly populations [33]. Future work should therefore explore more robust frameworks for cross-population generalization, or population-specific models tailored to these groups.

In addition, the wide SBP range of the Aurora-BP dataset increases the risk of train-test distribution shifts, which may also partly explain its suboptimal accuracy. Another contributing factor may be the normalization strategy. Specifically, PPG of the OPPO-BP dataset was normalized by that of the Aurora-BP dataset, which may not adequately represent the PPG distribution of the test set. This issue is particularly relevant because different wearable devices can exhibit varying PPG amplitude ranges.

### Limitations

Although this study proposed an effective framework to integrated multiple sources of features, we only utilized PPG signal as input. It is easy for wearable deployment but may suffer from the limitation of less informative features. For future work, new framework can be proposed to make full use of complementary information from multi-modal and multi-wavelength PPG signals, hopefully capture more about vascular dynamics to further enhance the performance.

## 6. Conclusion

This study introduced RACGBPNet, a novel deep learning framework for cuffless BP estimation and hypertension detection, which effectively and efficiently integrates handcrafted features, deep PPG representations, and demographic information. The model not only achieved superior estimation accuracy on a large public dataset but also effectively addressed class imbalance to improve hypertension detection. In long-term validation, RACGBPNet maintained strong compliance with AAMI standards under baseline calibration, demonstrating a practical balance between accuracy and usability in real-world monitoring. Nevertheless, the model exhibited reduced variability compared with reference BP. Future work should focus on enriching datasets with samples exhibiting larger deviations from individual baselines, advancing the robustness and clinical utility of wearable and cuffless BP monitoring systems.

## Data Availability

No data available in this study.

## Notes

### Competing Interest Statement

This study was partially sponsored by the Research Fund from OPPO Health Lab, Guangdong OPPO Mobile Telecommunications Corp., Ltd. The authors, Xiaoyu Li, Guangpu Zhu and Yelei Li, are the employees of the sponsoring company and were involved in the procedure of data acquisition of the OPPO-BP dataset. However, the design of the study, interpretation of the results, and preparation of the manuscript were carried out independently by the other authors. This division of roles was established to ensure full scientific objectivity and safeguard the integrity of the research process. All authors had unrestricted access to the data and retained full authority over the decision to submit the manuscript for publication.

### Funding Statement

Yes

### Author Declarations

The OPPO-BP dataset was collected in accordance with a standardized home BP monitoring protocol, which was approved by the Ethics Committee of the Tibet Branch of the West China Hospital of Sichuan University, and written informed consent was obtained from all participants prior to enrollment. The Microsoft Aurora BP dataset used in our study was requested from the Microsoft research team, which has been de-identified by them.

## References

1. Schutte AE, Kollias A, Stergiou GS. Blood pressure and its variability: classic and novel measurement techniques. Nature Reviews Cardiology. Schutte AE, Kollias A, Stergiou GS. Blood pressure and its variability: classic and novel measurement techniques. Nature Reviews Cardiology. 2022;19(10):643–54. 10.1038/s41569-022-00690-0

2. Zhou Y, Jia L, Lu B, Gu G, Hu H, Zhang Z, et al. Updated hypertension prevalence, awareness, and control rates based on the 2017ACC/AHA high blood pressure guideline. J Clin Hypertens (Greenwich). Zhou Y, Jia L, Lu B, Gu G, Hu H, Zhang Z, et al. Updated hypertension prevalence, awareness, and control rates based on the 2017ACC/AHA high blood pressure guideline. J Clin Hypertens (Greenwich). 2019;21(6):758-65. 10.1111/jch.13564 PMID:31131983

3. Green BB, Anderson ML, Campbell J, Cook AJ, Ehrlich K, Evers S, et al. Blood pressure checks and diagnosing hypertension (BP-CHECK): Design and methods of a randomized controlled diagnostic study comparing clinic, home, kiosk, and 24-hour ambulatory BP monitoring. Contemporary clinical trials. Green BB, Anderson ML, Campbell J, Cook AJ, Ehrlich K, Evers S, et al. Blood pressure checks and diagnosing hypertension (BP-CHECK): Design and methods of a randomized controlled diagnostic study comparing clinic, home, kiosk, and 24-hour ambulatory BP monitoring. Contemporary clinical trials. 2019;79:1-13.

4. Viera AJ, Hinderliter AL, Kshirsagar AV, Fine J, Dominik R. Reproducibility of masked hypertension in adults with untreated borderline office blood pressure: comparison of ambulatory and home monitoring. American journal of hypertension. Viera AJ, Hinderliter AL, Kshirsagar AV, Fine J, Dominik R. Reproducibility of masked hypertension in adults with untreated borderline office blood pressure: comparison of ambulatory and home monitoring. American journal of hypertension. 2010;23(11):1190–7.

5. Agarwal R, Light RP. The effect of measuring ambulatory blood pressure on nighttime sleep and daytime activity—implications for dipping. Clinical Journal of the American Society of Nephrology. Agarwal R, Light RP. The effect of measuring ambulatory blood pressure on nighttime sleep and daytime activity—implications for dipping. Clinical Journal of the American Society of Nephrology. 2010;5(2):281–5.

6. Schutte AE. Wearable cuffless blood pressure tracking: when will they be good enough? Journal of Human Hypertension. Schutte AE. Wearable cuffless blood pressure tracking: when will they be good enough? Journal of Human Hypertension. 2024;38(9):669–72.

7. Mukkamala R, Hahn J-O, Inan OT, Mestha LK, Kim C-S, Töreyin H, et al. Toward ubiquitous blood pressure monitoring via pulse transit time: theory and practice. IEEE transactions on biomedical engineering. Mukkamala R, Hahn J-O, Inan OT, Mestha LK, Kim C-S, Töreyin H, et al. Toward ubiquitous blood pressure monitoring via pulse transit time: theory and practice. IEEE transactions on biomedical engineering. 2015;62(8):1879–901.

8. Ding X-R, Zhang Y-T, editors. Photoplethysmogram intensity ratio: A potential indicator for improving the accuracy of PTT-based cuffless blood pressure estimation. 2015 37th annual international conference of the IEEE engineering in medicine and biology society (EMBC); 2015: IEEE.

9. Liu Q, Yan BP, Yu C-M, Zhang Y-T, Poon CC. Attenuation of systolic blood pressure and pulse transit time hysteresis during exercise and recovery in cardiovascular patients. IEEE Transactions on Biomedical Engineering. Liu Q, Yan BP, Yu C-M, Zhang Y-T, Poon CC. Attenuation of systolic blood pressure and pulse transit time hysteresis during exercise and recovery in cardiovascular patients. IEEE Transactions on Biomedical Engineering. 2013;61(2):346–52.

10. Ma Y, Choi J, Hourlier-Fargette A, Xue Y, Chung HU, Lee JY, et al. Relation between blood pressure and pulse wave velocity for human arteries. Proceedings of the National Academy of Sciences. Ma Y, Choi J, Hourlier-Fargette A, Xue Y, Chung HU, Lee JY, et al. Relation between blood pressure and pulse wave velocity for human arteries. Proceedings of the National Academy of Sciences. 2018;115(44):11144–9.

11. Pandit JA, Lores E, Batlle D. Cuffless blood pressure monitoring: promises and challenges. Clinical Journal of the American Society of Nephrology. Pandit JA, Lores E, Batlle D. Cuffless blood pressure monitoring: promises and challenges. Clinical Journal of the American Society of Nephrology. 2020;15(10):1531–8.

12. El-Hajj C, Kyriacou PA. A review of machine learning techniques in photoplethysmography for the non-invasive cuff-less measurement of blood pressure. Biomedical Signal Processing and Control. El-Hajj C, Kyriacou PA. A review of machine learning techniques in photoplethysmography for the non-invasive cuff-less measurement of blood pressure. Biomedical Signal Processing and Control. 2020;58:101870.

13. Senturk U, Polat K, Yucedag I. A non-invasive continuous cuffless blood pressure estimation using dynamic recurrent neural networks. Applied Acoustics. Senturk U, Polat K, Yucedag I. A non-invasive continuous cuffless blood pressure estimation using dynamic recurrent neural networks. Applied Acoustics. 2020;170:107534.

14. Liu Q, Zheng Y, Zhang Y, Poon CC. Beats-to-beats estimation of blood pressure during supine cycling exercise using a probabilistic nonparametric method. IEEE Access. Liu Q, Zheng Y, Zhang Y, Poon CC. Beats-to-beats estimation of blood pressure during supine cycling exercise using a probabilistic nonparametric method. IEEE Access. 2021;9:115655–63.

15. Hong J, Gao J, Liu Q, Zhang Y, Zheng Y, editors. Deep learning model with individualized fine-tuning for dynamic and beat-to-beat blood pressure estimation. 2021 IEEE 17th International Conference on Wearable and Implantable Body Sensor Networks (BSN); 2021: IEEE.

16. Wang W, Mohseni P, Kilgore KL, Najafizadeh L. Cuff-less blood pressure estimation from photoplethysmography via visibility graph and transfer learning. IEEE Journal of Biomedical and Health Informatics. Wang W, Mohseni P, Kilgore KL, Najafizadeh L. Cuff-less blood pressure estimation from photoplethysmography via visibility graph and transfer learning. IEEE Journal of Biomedical and Health Informatics. 2021;26(5):2075–85.

17. Su P, Ding X-R, Zhang Y-T, Liu J, Miao F, Zhao N, editors. Long-term blood pressure prediction with deep recurrent neural networks. 2018 IEEE EMBS International conference on biomedical & health informatics (BHI); 2018: IEEE.

18. Jeong DU, Lim KM. Combined deep CNN–LSTM network-based multitasking learning architecture for noninvasive continuous blood pressure estimation using difference in ECG-PPG features. Scientific Reports. Jeong DU, Lim KM. Combined deep CNN–LSTM network-based multitasking learning architecture for noninvasive continuous blood pressure estimation using difference in ECG-PPG features. Scientific Reports. 2021;11(1):13539.

19. Ma C, Zhang P, Song F, Sun Y, Fan G, Zhang T, et al. KD-informer: A cuff-less continuous blood pressure waveform estimation approach based on single photoplethysmography. IEEE Journal of Biomedical and Health Informatics. Ma C, Zhang P, Song F, Sun Y, Fan G, Zhang T, et al. KD-informer: A cuff-less continuous blood pressure waveform estimation approach based on single photoplethysmography. IEEE Journal of Biomedical and Health Informatics. 2022;27(5):2219–30.

20. Ma C, Zhang P, Song F, Liu Z, Feng Y, He Y, et al. UPR-BP: Unsupervised Photoplethysmography Representation Learning for Noninvasive Blood Pressure Estimation. IEEE Transactions on Artificial Intelligence. Ma C, Zhang P, Song F, Liu Z, Feng Y, He Y, et al. UPR-BP: Unsupervised Photoplethysmography Representation Learning for Noninvasive Blood Pressure Estimation. IEEE Transactions on Artificial Intelligence. 2024;5(9):4696–707.

21. Liu Z-D, Li Y, Zhang Y-T, Zeng J, Chen Z-X, Liu J-K, et al. HGCTNet: handcrafted feature-guided CNN and transformer network for wearable cuffless blood pressure measurement. IEEE Journal of Biomedical and Health Informatics. Liu Z-D, Li Y, Zhang Y-T, Zeng J, Chen Z-X, Liu J-K, et al. HGCTNet: handcrafted feature-guided CNN and transformer network for wearable cuffless blood pressure measurement. IEEE Journal of Biomedical and Health Informatics. 2024;28(7):3882–94.

22. Nour M, Polat K. Automatic classification of hypertension types based on personal features by machine learning algorithms. Mathematical Problems in Engineering. Nour M, Polat K. Automatic classification of hypertension types based on personal features by machine learning algorithms. Mathematical Problems in Engineering. 2020;2020(1):2742781.

23. Evdochim L, Dobrescu D, Halichidis S, Dobrescu L, Stanciu S. Hypertension detection based on photoplethysmography signal morphology and machine learning techniques. Applied Sciences. Evdochim L, Dobrescu D, Halichidis S, Dobrescu L, Stanciu S. Hypertension detection based on photoplethysmography signal morphology and machine learning techniques. Applied Sciences. 2022;12(16):8380.

24. Gupta K, Jiwani N, Afreen N, editors. Blood pressure detection using CNN-LSTM model. 2022 IEEE 11th International Conference on Communication Systems and Network Technologies (CSNT); 2022: IEEE.

25. Han M, Lee Y-R, Park T, Ihm S-H, Pyun WB, Burkard T, et al. Feasibility and measurement stability of smartwatch-based cuffless blood pressure monitoring: A real-world prospective observational study. Hypertension Research. Han M, Lee Y-R, Park T, Ihm S-H, Pyun WB, Burkard T, et al. Feasibility and measurement stability of smartwatch-based cuffless blood pressure monitoring: A real-world prospective observational study. Hypertension Research. 2023;46(4):922–31.

26. Miao F, Liu Z-D, Liu J-K, Wen B, He Q-Y, Li Y. Multi-sensor fusion approach for cuff-less blood pressure measurement. IEEE journal of biomedical and health informatics. Miao F, Liu Z-D, Liu J-K, Wen B, He Q-Y, Li Y. Multi-sensor fusion approach for cuff-less blood pressure measurement. IEEE journal of biomedical and health informatics. 2019;24(1):79–91.

27. Liu Z, Miao F, Wang R, Liu J, Wen B, Li Y, editors. Cuff-less blood pressure measurement based on deep convolutional neural network. 2019 41st Annual International Conference of the IEEE Engineering in Medicine and Biology Society (EMBC); 2019: IEEE.

28. Elgendi M, Galli V, Ahmadizadeh C, Menon C. Dataset of Psychological Scales and Physiological Signals Collected for Anxiety Assessment Using a Portable Device. Data [Internet]. 2022; 7(9).

29. Johnson AE, Pollard TJ, Shen L, Lehman L-wH, Feng M, Ghassemi M, et al. MIMIC-III, a freely accessible critical care database. Scientific data. Johnson AE, Pollard TJ, Shen L, Lehman L-wH, Feng M, Ghassemi M, et al. MIMIC-III, a freely accessible critical care database. Scientific data. 2016;3(1):1–9.

30. Lee H-C, Park Y, Yoon SB, Yang SM, Park D, Jung C-W. VitalDB, a high-fidelity multi-parameter vital signs database in surgical patients. Scientific Data. Lee H-C, Park Y, Yoon SB, Yang SM, Park D, Jung C-W. VitalDB, a high-fidelity multi-parameter vital signs database in surgical patients. Scientific Data. 2022;9(1):279.

31. Wang W, Mohseni P, Kilgore KL, Najafizadeh L. PulseDB: A large, cleaned dataset based on MIMIC-III and VitalDB for benchmarking cuff-less blood pressure estimation methods. Frontiers in Digital Health. Wang W, Mohseni P, Kilgore KL, Najafizadeh L. PulseDB: A large, cleaned dataset based on MIMIC-III and VitalDB for benchmarking cuff-less blood pressure estimation methods. Frontiers in Digital Health. 2023;4:1090854.

32. Mieloszyk R, Twede H, Lester J, Wander J, Basu S, Cohn G, et al. A comparison of wearable tonometry, photoplethysmography, and electrocardiography for cuffless measurement of blood pressure in an ambulatory setting. IEEE journal of biomedical and health informatics. Mieloszyk R, Twede H, Lester J, Wander J, Basu S, Cohn G, et al. A comparison of wearable tonometry, photoplethysmography, and electrocardiography for cuffless measurement of blood pressure in an ambulatory setting. IEEE journal of biomedical and health informatics. 2022;26(7):2864–75.

33. Liu Z-D, Li Y, Zhang Y-T, Zeng J, Chen Z-X, Cui Z-W, et al. Cuffless blood pressure measurement using smartwatches: a large-scale validation study. IEEE journal of biomedical and health informatics. Liu Z-D, Li Y, Zhang Y-T, Zeng J, Chen Z-X, Cui Z-W, et al. Cuffless blood pressure measurement using smartwatches: a large-scale validation study. IEEE journal of biomedical and health informatics. 2023;27(9):4216–27.

34. Poon C, Zhang Y, editors. Cuff-less and noninvasive measurements of arterial blood pressure by pulse transit time. 2005 IEEE engineering in medicine and biology 27th annual conference; 2006: IEEE.

35. Ding XR, Zhang YT, Liu J, Dai WX, Tsang HK. Continuous Cuffless Blood Pressure Estimation Using Pulse Transit Time and Photoplethysmogram Intensity Ratio. IEEE Transactions on Biomedical Engineering. Ding XR, Zhang YT, Liu J, Dai WX, Tsang HK. Continuous Cuffless Blood Pressure Estimation Using Pulse Transit Time and Photoplethysmogram Intensity Ratio. IEEE Transactions on Biomedical Engineering. 2016;63(5):964–72. 10.1109/TBME.2015.2480679

36. Liu J, Yan BP, Zhang YT, Ding XR, Su P, Zhao N. Multi-Wavelength Photoplethysmography Enabling Continuous Blood Pressure Measurement With Compact Wearable Electronics. IEEE Transactions on Biomedical Engineering. Liu J, Yan BP, Zhang YT, Ding XR, Su P, Zhao N. Multi-Wavelength Photoplethysmography Enabling Continuous Blood Pressure Measurement With Compact Wearable Electronics. IEEE Transactions on Biomedical Engineering. 2019;66(6):1514–25. 10.1109/TBME.2018.2874957

37. Qin C, Li Y, Liu C, Ma X. Cuff-Less Blood Pressure Prediction Based on Photoplethysmography and Modified ResNet. Bioengineering [Internet]. 2023; 10(4).

38. Zheng Y, Huang H, Gao J, Hong J, Wu S, Zhang Y, et al. UTransBPNet for cuffless and calibration-free blood pressure estimation under dynamic conditions. Scientific Reports. Zheng Y, Huang H, Gao J, Hong J, Wu S, Zhang Y, et al. UTransBPNet for cuffless and calibration-free blood pressure estimation under dynamic conditions. Scientific Reports. 2025;15(1):17654. 10.1038/s41598-025-02963-3

39. Sel K, Mohammadi A, Pettigrew RI, Jafari R. Physics-informed neural networks for modeling physiological time series for cuffless blood pressure estimation. npj Digital Medicine. Sel K, Mohammadi A, Pettigrew RI, Jafari R. Physics-informed neural networks for modeling physiological time series for cuffless blood pressure estimation. npj Digital Medicine. 2023;6(1):110. 10.1038/s41746-023-00853-4

40. Sola J, Arderiu A, Almeida TP, Fallet S, Yazdani S, Haddad S, et al. The quest for blood pressure markers in photoplethysmography and its applications in digital health. Frontiers in Digital Health. Sola J, Arderiu A, Almeida TP, Fallet S, Yazdani S, Haddad S, et al. The quest for blood pressure markers in photoplethysmography and its applications in digital health. Frontiers in Digital Health. 2025;Volume 7 - 2025. 10.3389/fdgth.2025.1518322

41. Chalmers J, MacMahon S, Mancia G, Whitworth J, Beilin L, Hansson L, et al. 1999 World Health Organization-International Society of Hypertension Guidelines for the management of hypertension. Guidelines sub-committee of the World Health Organization. Clinical and experimental hypertension (New York, NY: 1993). Chalmers J, MacMahon S, Mancia G, Whitworth J, Beilin L, Hansson L, et al. 1999 World Health Organization-International Society of Hypertension Guidelines for the management of hypertension. Guidelines sub-committee of the World Health Organization. Clinical and experimental hypertension (New York, NY: 1993). 1999;21(5-6):1009-60.

42. Zhang ZM, Chen S, Liang YZ. Baseline correction using adaptive iteratively reweighted penalized least squares. Analyst. Zhang ZM, Chen S, Liang YZ. Baseline correction using adaptive iteratively reweighted penalized least squares. Analyst. 2010;135(5):1138–46. 10.1039/b922045c PMID:20419267

43. Cisnal A, Li Y, Fuchs B, Ejtehadi M, Riener R, Paez-Granados D. Robust Feature Selection for BP Estimation in Multiple Populations: Towards Cuffless Ambulatory BP Monitoring. IEEE Journal of Biomedical and Health Informatics. Cisnal A, Li Y, Fuchs B, Ejtehadi M, Riener R, Paez-Granados D. Robust Feature Selection for BP Estimation in Multiple Populations: Towards Cuffless Ambulatory BP Monitoring. IEEE Journal of Biomedical and Health Informatics. 2024;28(10):5768–79. 10.1109/JBHI.2024.3411693

44. Cho K, van Merrienboer B, Gulcehre C, Bahdanau D, Bougares F, Schwenk H, et al. Learning Phrase Representations using RNN Encoder-Decoder for Statistical Machine Translation 2014 June 01, 2014:[arXiv:1406.078 p.]. Available from: https://ui.adsabs.harvard.edu/abs/2014arXiv1406.1078C.

45. Balntas V, Riba E, Ponsa D, Mikolajczyk K, editors. Learning local feature descriptors with triplets and shallow convolutional neural networks. Bmvc; 2016.

46. Miao F, Wen B, Hu Z, Fortino G, Wang X-P, Liu Z-D, et al. Continuous blood pressure measurement from one-channel electrocardiogram signal using deep-learning techniques. Artificial Intelligence in Medicine. Miao F, Wen B, Hu Z, Fortino G, Wang X-P, Liu Z-D, et al. Continuous blood pressure measurement from one-channel electrocardiogram signal using deep-learning techniques. Artificial Intelligence in Medicine. 2020;108:101919. 10.1016/j.artmed.2020.101919

